# Development of Self-Assessment Tools for Osteoporosis among Postmenopausal Vietnamese Women: A Machine Learning Approach

**DOI:** 10.1101/2025.10.27.25338918

**Authors:** Tung-Lam Nguyen, Thuy Trang Nguyen, Christine Pallota, Long Q. Khuong, My Hanh Bui

**Affiliations:** Pleasantville High School, NY, USA; Hanoi Medical University, Hanoi Vietnam; University of Pennsylvania, PA, USA; Hanoi Medical University Hospital, Hanoi Vietnam

**Author notes:** **Correspondence:** My Hanh Bui, MD, PhD, Hanoi Medical University & Hanoi Medical University Hospital, 1 Ton That Tung, Hanoi, Vietnam. **Fundings:** None. **Author Contributions:** Conceptualization: TLN, TTN, and MHB. Formal analysis: TLN. Methodology: TLN and LQK. Visualization: TLN. Writing–original draft: TLN. Writing–review & editing: TLN, TTN, CP, LQK, MHB. All authors approved the final version of the manuscript.

## Abstract

**Background:** Osteoporosis is a major health concern in Vietnam due to a rise in aging rates. However, cost-effective early screening tools tailored to the Vietnamese population are lacking. In this study, we applied a machine learning approach to develop self-assessment tools for osteoporosis in menopausal women.

**Methods:** We used retrospective data from 15,670 postmenopausal Vietnamese women extracted from electronic medical records. Bone mineral density (BMD) measurements of the lumbar vertebrae (L1–L4) and the left and right femoral necks were obtained using the dual-energy X-ray absorptiometry (DXA) system. Osteoporosis was defined as a BMD T-score of < -2.5. Eight algorithms (Logistic Regression, Decision Trees, Random Forest, XGBoost, K-Nearest Neighbors, Neural Networks, Lasso and Ridge regressions, and Naïve Bayes) were utilized to develop prediction algorithms for each anatomical site. The main predictors included age, menopause age, weight, height, lifestyles, medical history and comorbidities. Cross-validation was employed to prevent overfitting.

**Results:** The prevalence of osteoporosis, as determined by BMD, varied across anatomical sites, ranging from 22% at the left and right femoral necks to 18.3–35.5% across the lumbar vertebrae. The performance of the models varied slightly among algorithms, although the differences were not substantial. Considering the balance between performance and simplicity, logistic regression was selected as the final algorithm. The final models were developed using four predictors: age, menopause age, height, and BMI, with the area under the curve (AUC) ranging from 0.75 at lumbar vertebrae L4 to 0.84 at right femur neck. Our models demonstrated superior performance compared to existing tools, such as OSTA, which were developed for the general Asian population.

**Conclusion:** The newly developed self-assessment tools were shown to be simple and effective in predicting osteoporosis among postmenopausal Vietnamese women. These tools have the potential to serve as early screening instruments for osteoporosis, particularly in resource-limited settings.

## Introduction

Osteoporosis is a metabolic disorder characterized by the weakening of the bone caused by an altered bone microstructure [1]. It has become a global health concern, affecting an estimated 18.3% of the world’s population, with prevalence rates reaching 23.1% in women and 11.7% in men between 2021 to 2022 [2]. The incidence is growing rapidly, projected to reach 263.2 million people between 2030 and 2024 as in comparison to 2019 with 41.5 million cases [3]. Worldwide, osteoporosis-related fractures cost hundreds of billions of dollars annually which greatly contribute to economic burden for low-income countries. Women are particularly more susceptible to osteoporosis than man during post-menopause stage. As global population ages, the incidence and impact of hip fractures will increase sharply making it a critical focus for public health intervention.

Vietnam is a low-middle income country with a population exceeding 100 million, with a significant portion of the population being within the older age brackets. In 2024, individuals of 65 or older accounts for about 8% of the Vietnamese population. It is projected to reach approximately 14% by 2035 and increase to 20.4% by 2050 [4]. Osteoporosis has been an emerging concern in Vietnam, particularly among postmenopausal women. In a recent study of Vietnam, the prevalence of osteoporosis in women above the age of 50 was about 29% — a prevalence higher than the global average [5]. These Vietnamese individuals have a high likelihood of osteoporosis due to numerous factors: lack of calcium intake in traditional Vietnamese diet, sedentary lifestyles from rapid urbanization, and genetic factors that influence peak bone mass. While DXA is considered the gold standard for bone density measurement, it requires advanced technology, trained technicians, and substantial financial resources to operate, which presents a significant barrier for many individuals in Vietnam. Therefore, a scalable and cost-effective approach to osteoporosis screening is essential. Self-assessment tools provide a proactive solution by empowering individuals to evaluate their risk and seek medical advice when necessary. Requiring fewer resources, they can be widely distributed, making them accessible to a larger segment of the population.

The Osteoporosis Self-Assessment Tool (OST) is one of the oldest and simplest ways to identify people at risk of osteoporosis using a statistical approach developed by Koh. et al in 2001 [6]. This tool can identify the population that are likely to have osteoporosis in men and women. More complex tools have then been developed with additional factors and characteristics to improve performance of prediction. This includes OSTA, FRAX, ORAI, SCORE, ORISIS, ABONE, and MOST. However, it was observed that the performance of osteoporosis self-assessment tools can vary depending on the demographic. For instance, Bui et al.’s study, which evaluated in various anatomical sites the performance of OSTA, for Asian, compared to OSTC, for Chinese population. It was found that OSTA and OSTC performance greatly differ [7]. This finding highlights the impact of demographic on model performance and underscores the need for tools to be calibrated to specific populations [7]. By identifying individuals at highest risk with readily available inputs like age, weight, or basic clinical data from local data, predictive models will help healthcare providers focus on limited resources where they are needed most. This prevents unnecessary testing and optimizes early intervention. In communities with limited access to advanced diagnostics, models calibrated with local data can guide preventive measures, all while relying on low-cost tools.

This study was conducted to develop a prediction model for osteoporosis among postmenopausal women in Vietnam using machine learning techniques. The development of this model can be beneficial to health care organizations in many ways: to target and prioritize interventions, act as an assistant tool to enhance clinical decision-making and prescreening tools for high-risk patients.

## Methodology

### Data Source and Study Population

This research used a retrospective dataset of women who visited the Department of Functional Exploration at Hanoi Medical University Hospital, Vietnam, extracted from the Health Information System (HIS) in during 2021 and 2023 with fully anonymized data extraction procedures. The selection criteria were: (1) postmenopausal women, aged > 40 years, (2) without any anatomical abnormalities on the anatomical sites being investigated, and (3) who had not undergone spinal or femur surgery. The final dataset after the extraction was 15,670 women. As the data were de-identified, no personally identifiable information was included, ensuring that participants could not be re-identified or traced. As a result, the study posed no risk to participant’s privacy, confidentiality, or well-being. No direct interaction with participants occurred, and no intervention or data collection was performed specifically for this study. The data are part of an on-going project that received ethical approval from the Institutional Review Board of Hanoi Medical University (IRB number: 00003121). The study adhered to ethical principles outlined in the Declaration of Helsinki, ensuring the responsible and ethical use of secondary data.

### Outcome definition

The outcome of interest was osteoporosis (yes/no), defined based on bone mineral density (BMD), measured using a DXA system. The BMD values were converted to T-scores, representing the number of standard deviations from the mean peak bone mass. According to the World Health Organization (WHO) criteria, a T-score ≤ –2.5 indicates the presence of osteoporosis [17]. We evaluated osteoporosis at six anatomical sites: the left femur, right femur, lumbar vertebrae L1, L2, L3, and L4, as well as at any site overall.

### Candidate predictors

Predictors were selected based on domain knowledge and a review of the literature on existing self-assessment tools. Given the focus on screening, we considered two categories of predictors: (1) basic anthropometric measures, including age, height, weight, and body mass index (BMI), and (2) clinical characteristics, including history of bone fracture, family history of bone fracture, glucocorticoid use within the previous three months, rheumatoid disease, type 1 diabetes, thyroid disease, sexual dysfunction, chronic undernutrition, and alcohol consumption of at least three units in the past month. BMI was derived as weight (kg)/height^2^ (m). Participant characteristics were collected at the time of registration by specialist physicians

### Model Development

The general model-building strategy is illustrated in **Figure 1**. The dataset was randomly split into training and testing sets in an 80:20 ratio, with 80% of the original dataset (12,535 subjects) used for training and the remaining 20% for testing. For models requiring hyperparameter tuning, 10-fold cross-validation was conducted within the training dataset to select the optimal hyperparameters. The model-building strategy is repeated for all 8 algorithms, each with 7 anatomical sites.

**Figure 1.**
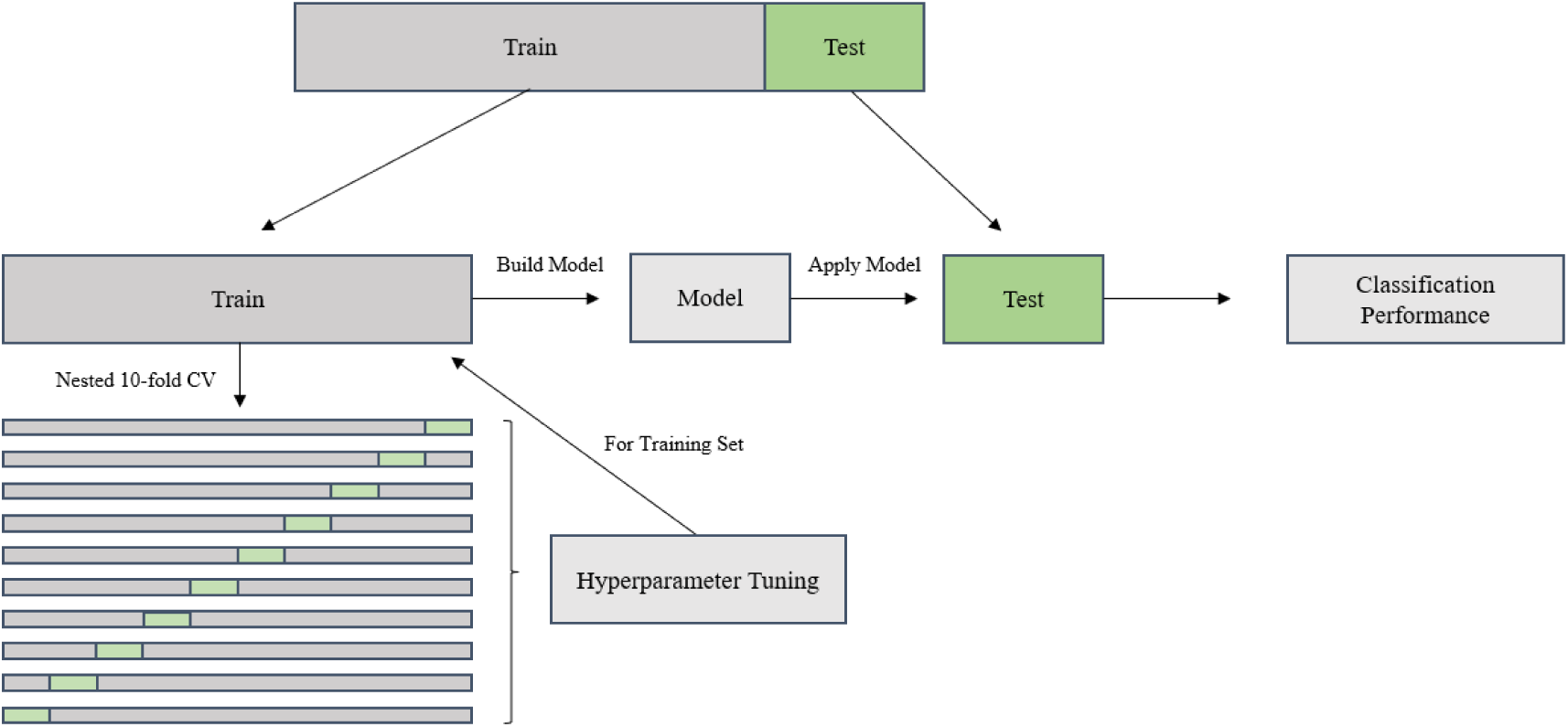
Flowchart of model training, parameter tuning, and performance evaluation.

### Handling Missing Values and Variable Selection

Outlier detection using z-scores was applied to identify anomalies in the data that could potentially distort the results. Values falling outside of ±3 standard deviations were considered outliers. The total proportion of missing values was 5%. Missing data were addressed using single imputation.

Mean Decrease Gini (MDG) in Random Forests was used to assess the importance of the features for feature selection. Random Forest was selected because MDG is a built-in metric in the algorithm. A high MDG score would indicate that a feature plays a significant role in splitting data [16]. Any features that have a significantly lower score were not included as the models’ predictors. Feature encoding was not necessary as there were no categorical variables after feature selection. Although weight had a high importance, it was not included as BMI provided weight information.

### Model Estimation

In the model development process, eight different machine learning algorithms were used and compared. The selection models were based on diversifying the choice of methodologies and the level of model complexity: (1) Logistic Regression (LR) can predict the probabilities of an event using the sigmoid function. It is known for its simplicity and interpretability which allows the model to provide interpretable coefficients, making it useful in healthcare fields [9]; (2) Decision Tree (DT) is a flowchart-like model with the dataset split into smaller subsets based on the feature values. The internal node represents the feature, and each branch of the decision tree represents a decision rule. When a criterion is met, the model reaches the leaf node, where the class label of the datapoint is determined. They are flexible and can naturally handle both categorical and numerical data [10]; (3) Random Forest (RF) is an ensemble method that creates multiple decision trees using different subsets of the data and different subsets of features. For classification, the final output is determined by averaging majority voting from all the individual trees. This helps in reducing overfitting and improving accuracy [10]; (4) LASSO involves a penalty on the absolute size of the coefficients, which can result in some coefficients being exactly zero. This effectively performs variable selection and regularization simultaneously [11]; (5) XGBoost (XGB) builds an ensemble of decision trees in a sequential manner, where each tree corrects the errors made by the previous ones. It uses a gradient descent algorithm to minimize a loss function and improve the model’s accuracy. It is known for its speed and performance, especially with large datasets [12]; (6) Neural networks (NN) have layers of interconnected nodes, or neurons which work together to recognize patterns in data. Neural networks can have multiple layers to capture more complex patterns. They learn through backpropagation, where they adjust the weights of the connections based on the error in the output compared to the expected result [13]; (7) Naïve Bayes (NB) is a family of probabilistic algorithms with Bayes’ theorem as its foundation. Bayes’ Theorem describes the probability of an event based on other information that might be relevant, essentially updating the estimates based on additional knowledge [14]; (8) K-nearest neighbors (KNN) examine the labels of a chosen number of data points surrounding a target data point to make predictions about the class that the data point falls into. It stores all available cases and classifies new cases based on a similarity measure. The object is assigned to the class most common among its K nearest neighbors. It is a simple and effective algorithm for classification and regression [15].

Cross validation was performed across all 8 algorithms through 10-fold cross validation for model evaluation and to tune the parameters. In this method, the training dataset is randomly divided into 10 approximate equal-sized folds. For each of the 10 repetitions, 9 of the fold is used as the training set and 1-fold is used as the testing set. Only the training set was used to preserve the integrity of the test set. The parameters are fine-tuned by grid search, ensuring the model performance is more realistic. The grid search uses a specified set of values for each hyperparameter, and the algorithms test all the possible combinations of hyperparameters. Logistic Regression did not require any grid search.

### Model Evaluation

Model testing was conducted across all sites for each algorithm using the corresponding optimized hyperparameters. All performance metrics were calculated on the testing dataset.

#### Discrimination

Discrimination was quantified with the area under the Receiver Operating Characteristic (ROC) curve (AUC) and a confusion matrix. AUC reflects how well the model ranks positive cases ahead of negatives: values close to 1 indicate better separation, whereas 0.5 means no better than random guessing. An AUC < 70% indicates poor classification, AUC between 70% and 80% indicates moderate classification, and AUC > 80% or higher reflects good classification [18].

The confusion matrix is built at a chosen probability cut-off, allowing computation of accuracy, sensitivity, and specificity. The optimal cut-off points for estimating sensitivity and specificity were determined using the Youden Index, defined as (sensitivity + specificity – 1) [19]. The confusion matrix consists of four categories: true positives (TP), false negatives (FN), false positives (FP), and true negatives (TN). From these quantities, we derived accuracy, sensitivity (true positive rate), and specificity (true negative rate) using the following formulas:

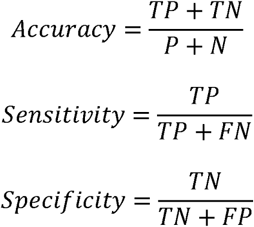

#### Calibration

To assess the reliability of predicted probabilities, calibration analysis was performed across all binary outcome models. Calibration curves were constructed by stratifying cases into deciles of predicted risk and plotting the mean predicted probability in each bin against the observed event rate. A well-calibrated model produces points that lie close to the 45-degree line, indicating alignment between predicted probabilities and actual outcomes [20]

## Results

**Table 1** compares the characteristics of participants in the training (N = 12,535) and testing (N = 3,135) datasets. The distribution of participants’ characteristics was similar between the two datasets. Participants in both sets had a mean age of 62 years. The average height was 151.8 cm in the training set and 151.9 cm in the testing set, while the mean weight was 53 kg in both groups, resulting in nearly identical mean BMI values (22.90 kg/m² in the training data and 22.88 kg/m² in the testing data). The mean age at menopause was approximately 49.0 years in both datasets.

**Table 1.** Participants characteristics by training and testing data.

| Characteristics | Training Data<br>N = 12,535 | Testing Data<br>N = 3,135 | p-value |
| --- | --- | --- | --- |
| Age, year – mean ± SD | 62.0 ± 10.0 | 62.0 ± 10.0 | 0.4 |
| Height, cm – mean ± SD | 151.8 ± 5.7 | 151.9 ± 5.6 | 0.4 |
| Weight, kg – mean ± SD | 53.0 ± 7.0 | 53.0 ± 7.0 | 0.8 |
| BMI, kg/m<sup>2</sup> – mean ± SD | 22.90 ± 2.8 | 22.88 ± 2.8 | 0.8 |
| Menopausal age – mean ± SD | 49.0 ± 3.9 | 49.2 ± 4.0 | 0.1 |
| History of bone fracture, % | 1,041 (8.3%) | 278 (8.9%) | 0.3 |
| Family history of bone fracture, % | 195 (1.6%) | 50 (1.6%) | 0.9 |
| Glucocorticoid within 3 months, % | 143 (1.1%) | 33 (1.1%) | 0.7 |
| Rheumatoid, % | 544 (4.3%) | 127 (4.1%) | 0.5 |
| Diabetes_type1, % | 38 (0.3%) | 5 (0.2%) | 0.2 |
| Thyroid disease, % | 1,239 (9.9%) | 327 (10%) | 0.4 |
| Sexual dysfunction, % | 2 (<0.1%) | 3 (<0.1%) | 0.1 |
| Chronic undernutrition, % | 5 (<0.1%) | 0 (0%) | 0.6 |
| Menopause age less than 46 years, % | 1,922 (15%) | 467 (15%) | 0.5 |
| Consuming alcohol more than 3 units, % | 5 (<0.1%) | 3 (<0.1%) | 0.2 |

SD: Standard Deviation

**Figure 2** presents the proportions of pre-osteoporosis and osteoporosis based on BMD classification. The prevalence of osteoporosis varied across anatomical sites, ranging from about 22% at the femoral necks to 35.5% at the third lumbar vertebra. Overall, 22.7% of participants were classified as osteoporosis at any anatomical site.

**Figure 2.**
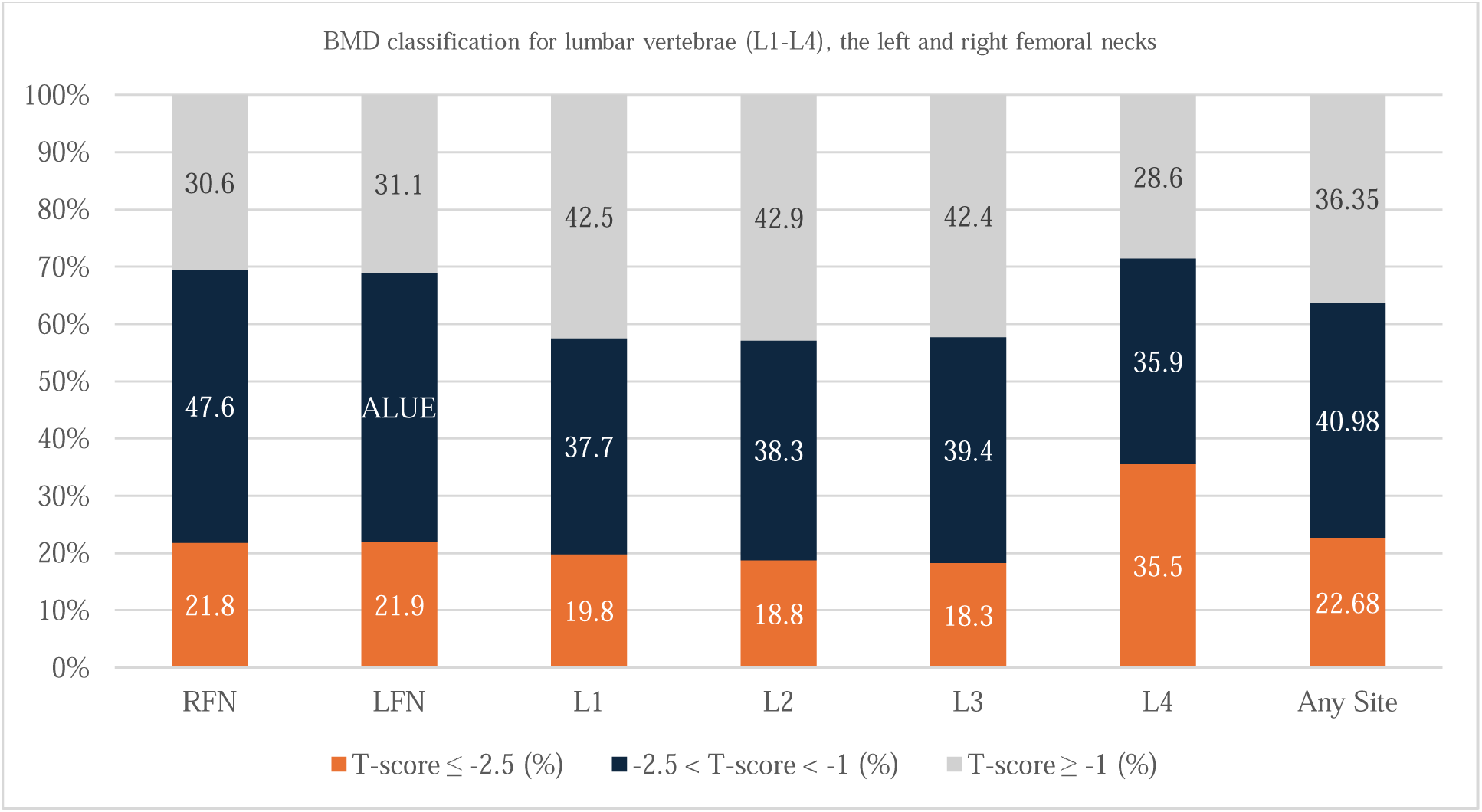
BMD classification of analytical sample. FN: femoral neck, LS1: lumbar vertebrae L1, LS2: lumbar vertebrae L2, LS3: lumbar vertebrae L3, LS4: lumbar vertebrae L4, LS: lumbar spine

**Figure 3** presents the feature importance based on the Mean Decrease Gini (MDG). Age, height, weight, and BMI demonstrated the highest MDG values across all anatomical sites, with age being the most important predictor, particularly for the lumbar vertebra. Family history of bone fracture, rheumatoid disease, thyroid disease, and menopause before age 46 showed relatively low importance, while all other predictors contributed little to none. Considering both the feature importance ranking and the trade-off between model performance and parsimony, four variables were selected for the main models: age, weight, BMI, and menopausal age.

**Figure 3.**
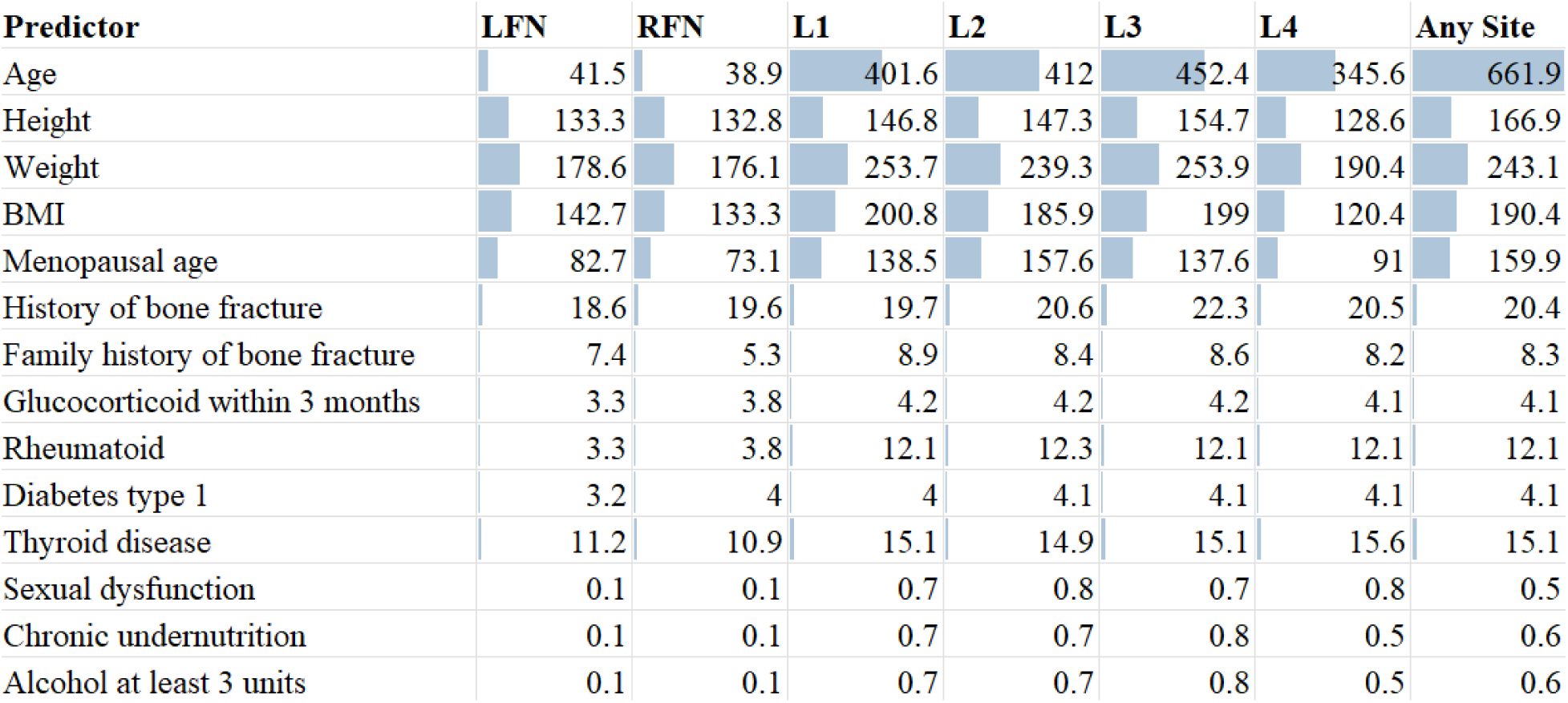
Importance of all predictors with Mean Decrease Gini.

**Table 2** and **Figure 4** compare the discrimination performance measures of eight algorithms. LR, LASSO, XGB, and NN demonstrated strong model performance with high AUC values across the sites (AUC 0.75–0.84). DT, RF, and NB showed moderate performance (AUC 0.73–0.78), while KNN performed the worst across most metrics (AUC < 0.73).

**Figure 4.**
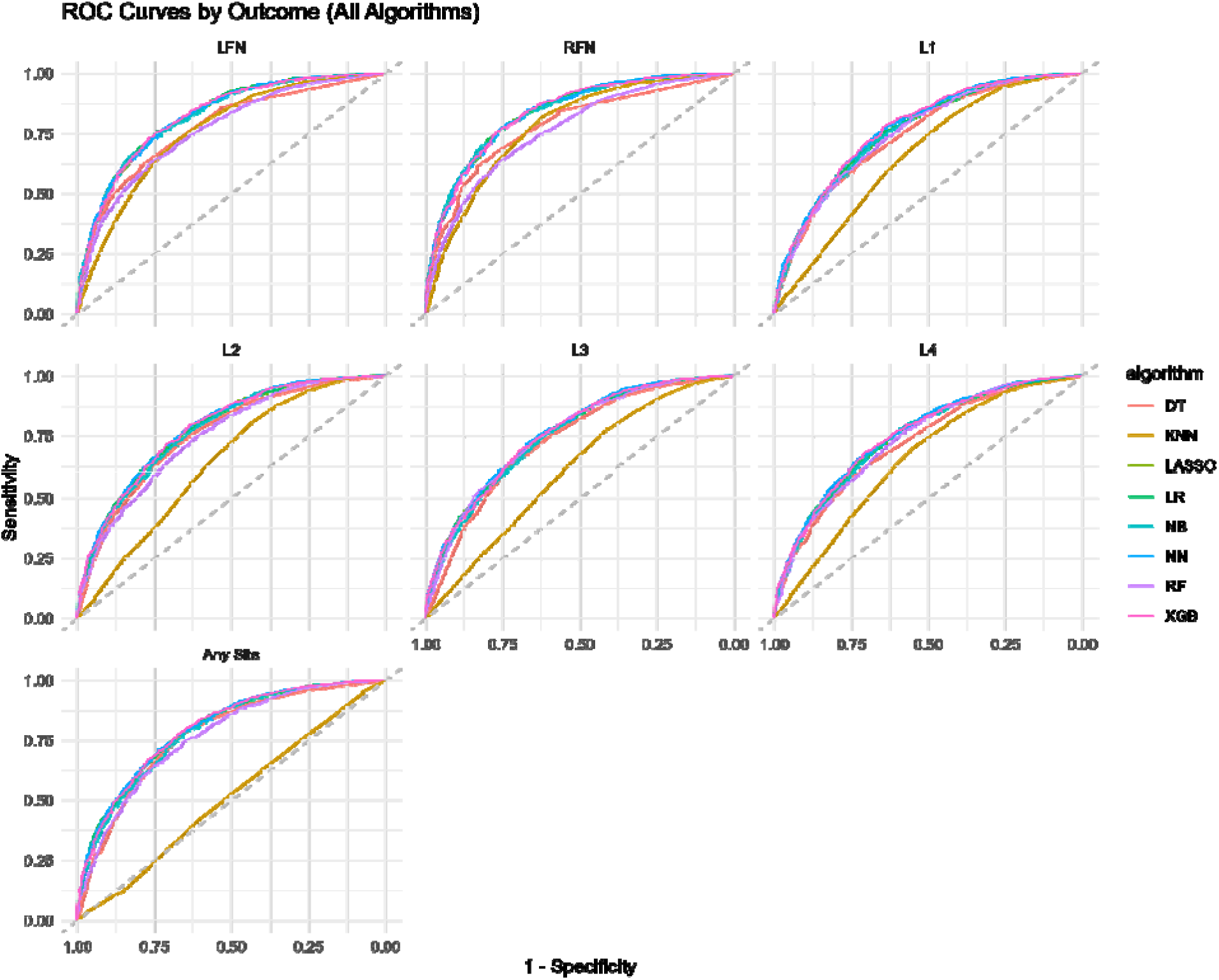
ROC curves for eight algorithms at all anatomical. sites FN: femoral neck, LS1: lumbar vertebrae L1, LS2: lumbar vertebrae L2, LS3: lumbar vertebrae L3, LS4: lumbar vertebrae L4. DT: Decision Tree, KNN: K-nearest Neighbors, LR: Logistic Regression, NB: Naïve Bayes, NN: Neural Networks, RF: Random Forest, XGB: XGBoost

**Table 2:** Model’s performance using testing data.

| Model | AUC (95% CI) | Cutoff | Accuracy | Sensitivity | Specificity | PPV | NPV |
| --- | --- | --- | --- | --- | --- | --- | --- |
| **Logistic Regression** |  |  |  |  |  |  |  |
| Left femoral neck | 0.82 (0.81-0.82) | 0.20 | 0.74 | 0.76 | 0.74 | 0.43 | 0.92 |
| Right femoral neck | 0.84 (0.82-0.84) | 0.17 | 0.74 | 0.80 | 0.73 | 0.41 | 0.94 |
| L1 | 0.76 (0.74-0.76) | 0.32 | 0.68 | 0.76 | 0.64 | 0.56 | 0.81 |
| L2 | 0.78 (0.77-0.78) | 0.36 | 0.70 | 0.73 | 0.69 | 0.59 | 0.81 |
| L3 | 0.76 (0.74-0.76) | 0.38 | 0.68 | 0.73 | 0.65 | 0.60 | 0.77 |
| L4 | 0.75 (0.73-0.75) | 0.33 | 0.68 | 0.69 | 0.68 | 0.53 | 0.81 |
| Any site | 0.80 (0.78-0.80) | 0.51 | 0.72 | 0.74 | 0.70 | 0.75 | 0.70 |
| **Decision Tree** |  |  |  |  |  |  |  |
| Left femoral neck | 0.77 (0.75-0.79) | 0.19 | 0.76 | 0.61 | 0.80 | 0.44 | 0.89 |
| Right femoral neck | 0.78 (0.76-0.80) | 0.25 | 0.79 | 0.61 | 0.83 | 0.47 | 0.90 |
| L1 | 0.74 (0.73-0.76) | 0.34 | 0.69 | 0.61 | 0.73 | 0.58 | 0.76 |
| L2 | 0.76 (0.74-0.78) | 0.33 | 0.69 | 0.71 | 0.68 | 0.57 | 0.80 |
| L3 | 0.73 (0.72-0.75) | 0.42 | 0.68 | 0.63 | 0.72 | 0.62 | 0.73 |
| L4 | 0.73 (0.71-0.74) | 0.32 | 0.70 | 0.58 | 0.76 | 0.56 | 0.77 |
| Any site | 0.77 (0.75-0.79) | 0.48 | 0.72 | 0.77 | 0.66 | 0.73 | 0.71 |
| **Random Forest** |  |  |  |  |  |  |  |
| Left femoral neck | 0.76 (0.75-0.76) | 0.16 | 0.68 | 0.80 | 0.65 | 0.37 | 0.93 |
| Right femoral neck | 0.76 (0.75-0.76) | 0.14 | 0.68 | 0.84 | 0.64 | 0.36 | 0.94 |
| L1 | 0.75 (0.74-0.75) | 0.26 | 0.62 | 0.82 | 0.51 | 0.50 | 0.82 |
| L2 | 0.75 (0.73-0.75) | 0.31 | 0.65 | 0.79 | 0.56 | 0.52 | 0.81 |
| L3 | 0.76 (0.74-0.76) | 0.36 | 0.65 | 0.74 | 0.59 | 0.57 | 0.76 |
| L4 | 0.74 (0.72-0.74) | 0.24 | 0.59 | 0.80 | 0.48 | 0.45 | 0.82 |
| Any site | 0.77 (0.75-0.77) | 0.51 | 0.70 | 0.75 | 0.65 | 0.72 | 0.68 |
| **LASSO** |  |  |  |  |  |  |  |
| Left femoral neck | 0.82 (0.81-0.82) | 0.21 | 0.75 | 0.76 | 0.74 | 0.43 | 0.92 |
| Right femoral neck | 0.84 (0.82-0.84) | 0.18 | 0.75 | 0.78 | 0.74 | 0.42 | 0.93 |
| L1 | 0.76 (0.74-0.76) | 0.33 | 0.68 | 0.75 | 0.64 | 0.56 | 0.81 |
| L2 | 0.78 (0.77-0.78) | 0.33 | 0.69 | 0.78 | 0.63 | 0.56 | 0.83 |
| L3 | 0.76 (0.74-0.76) | 0.37 | 0.68 | 0.74 | 0.64 | 0.60 | 0.77 |
| L4 | 0.75 (0.73-0.75) | 0.33 | 0.68 | 0.70 | 0.67 | 0.53 | 0.81 |
| Any site | 0.80 (0.78-0.80) | 0.50 | 0.72 | 0.74 | 0.70 | 0.75 | 0.70 |
| **XGBoost** |  |  |  |  |  |  |  |
| Left femoral neck | 0.82 (0.81-0.82) | 0.24 | 0.76 | 0.73 | 0.76 | 0.45 | 0.92 |
| Right femoral neck | 0.84 (0.82-0.84) | 0.22 | 0.77 | 0.75 | 0.77 | 0.44 | 0.93 |
| L1 | 0.77 (0.76-0.77) | 0.35 | 0.69 | 0.78 | 0.64 | 0.57 | 0.83 |
| L2 | 0.79 (0.77-0.79) | 0.35 | 0.70 | 0.79 | 0.64 | 0.57 | 0.83 |
| L3 | 0.76 (0.75-0.76) | 0.43 | 0.69 | 0.70 | 0.69 | 0.62 | 0.76 |
| L4 | 0.75 (0.74-0.75) | 0.36 | 0.70 | 0.68 | 0.71 | 0.55 | 0.81 |
| Any site | 0.80 (0.78-0.80) | 0.52 | 0.73 | 0.77 | 0.67 | 0.74 | 0.71 |
| **Neural Network** |  |  |  |  |  |  |  |
| Left femoral neck | 0.82 (0.81-0.82) | 0.22 | 0.74 | 0.76 | 0.73 | 0.43 | 0.92 |
| Right femoral neck | 0.84 (0.82-0.84) | 0.23 | 0.77 | 0.75 | 0.77 | 0.44 | 0.93 |
| L1 | 0.77 (0.76-0.77) | 0.36 | 0.69 | 0.78 | 0.64 | 0.57 | 0.82 |
| L2 | 0.79 (0.77-0.79) | 0.39 | 0.70 | 0.74 | 0.68 | 0.58 | 0.81 |
| L3 | 0.77 (0.75-0.77) | 0.42 | 0.69 | 0.74 | 0.65 | 0.61 | 0.77 |
| L4 | 0.76 (0.74-0.76) | 0.37 | 0.69 | 0.70 | 0.68 | 0.54 | 0.81 |
| Any site | 0.80 (0.78-0.80) | 0.57 | 0.72 | 0.72 | 0.73 | 0.76 | 0.69 |
| **Naïve Bayes** |  |  |  |  |  |  |  |
| Left femoral neck | 0.82 (0.80-0.82) | 0.27 | 0.79 | 0.67 | 0.82 | 0.49 | 0.90 |
| Right femoral neck | 0.84 (0.82-0.84) | 0.18 | 0.76 | 0.77 | 0.75 | 0.43 | 0.93 |
| L1 | 0.76 (0.75-0.76) | 0.34 | 0.69 | 0.72 | 0.66 | 0.57 | 0.80 |
| L2 | 0.78 (0.76-0.78) | 0.30 | 0.68 | 0.79 | 0.61 | 0.55 | 0.83 |
| L3 | 0.75 (0.74-0.75) | 0.34 | 0.67 | 0.75 | 0.61 | 0.59 | 0.77 |
| L4 | 0.75 (0.73-0.75) | 0.30 | 0.67 | 0.74 | 0.64 | 0.52 | 0.82 |
| Any site | 0.79 (0.77-0.79) | 0.45 | 0.72 | 0.78 | 0.65 | 0.73 | 0.71 |
| **K-Nearest Neighbor** |  |  |  |  |  |  |  |
| Left femoral neck | 0.76 (0.74-0.76) | 0.80 | 0.32 | 0.27 | 0.33 | 0.09 | 0.63 |
| Right femoral neck | 0.78 (0.76-0.78) | 0.85 | 0.34 | 0.18 | 0.38 | 0.06 | 0.66 |
| L1 | 0.66 (0.64-0.66) | 0.80 | 0.42 | 0.18 | 0.57 | 0.20 | 0.53 |
| L2 | 0.65 (0.63-0.65) | 0.78 | 0.42 | 0.20 | 0.56 | 0.21 | 0.54 |
| L3 | 0.62 (0.60-0.62) | 0.76 | 0.43 | 0.22 | 0.59 | 0.28 | 0.51 |
| L4 | 0.66 (0.64-0.66) | 0.74 | 0.40 | 0.29 | 0.46 | 0.22 | 0.55 |
| Any site | 0.51 (0.49-0.51) | 0.66 | 0.52 | 0.63 | 0.40 | 0.56 | 0.47 |

Sensitivity varied across models and anatomical sites. RF demonstrated the highest sensitivity overall, reaching 0.84 at the right femoral neck and maintaining strong performance across the lumbar sites (0.74–0.82). LR, LASSO, XGB, and NN showed consistently high sensitivity (0.69–0.80). NB performed moderately well (0.67–0.79), DT showed lower sensitivity (0.58–0.71), and KNN had the weakest performance, with sensitivity values as low as 0.18 (**Table 2**). LR, LASSO, XGB, NB, and NN consistently demonstrated high specificity, typically ranging from 0.74 to 0.76 across all sites. DT also showed strong specificity, particularly at the femoral neck sites (0.83–0.84), though its sensitivity was notably lower. RF exhibited lower specificity (0.48–0.65), and KNN had the lowest specificity overall (0.33–0.59) (**Table 2**).

**Figure 5** presents calibration plots for eight algorithms across seven anatomical sites. Across the anatomical sites, LR, LASSO, NB, NN, and RF demonstrated good calibration, with close alignment between predicted probabilities and observed proportions. In contrast, DT and XGB underperformed, with DT showing deviation from the diagonal line in the highest decile at the lumbar vertebrae, while XGB exhibited consistent deviation from the diagonal, indicating systematic overestimation of osteoporosis risk.

**Figure 5.**
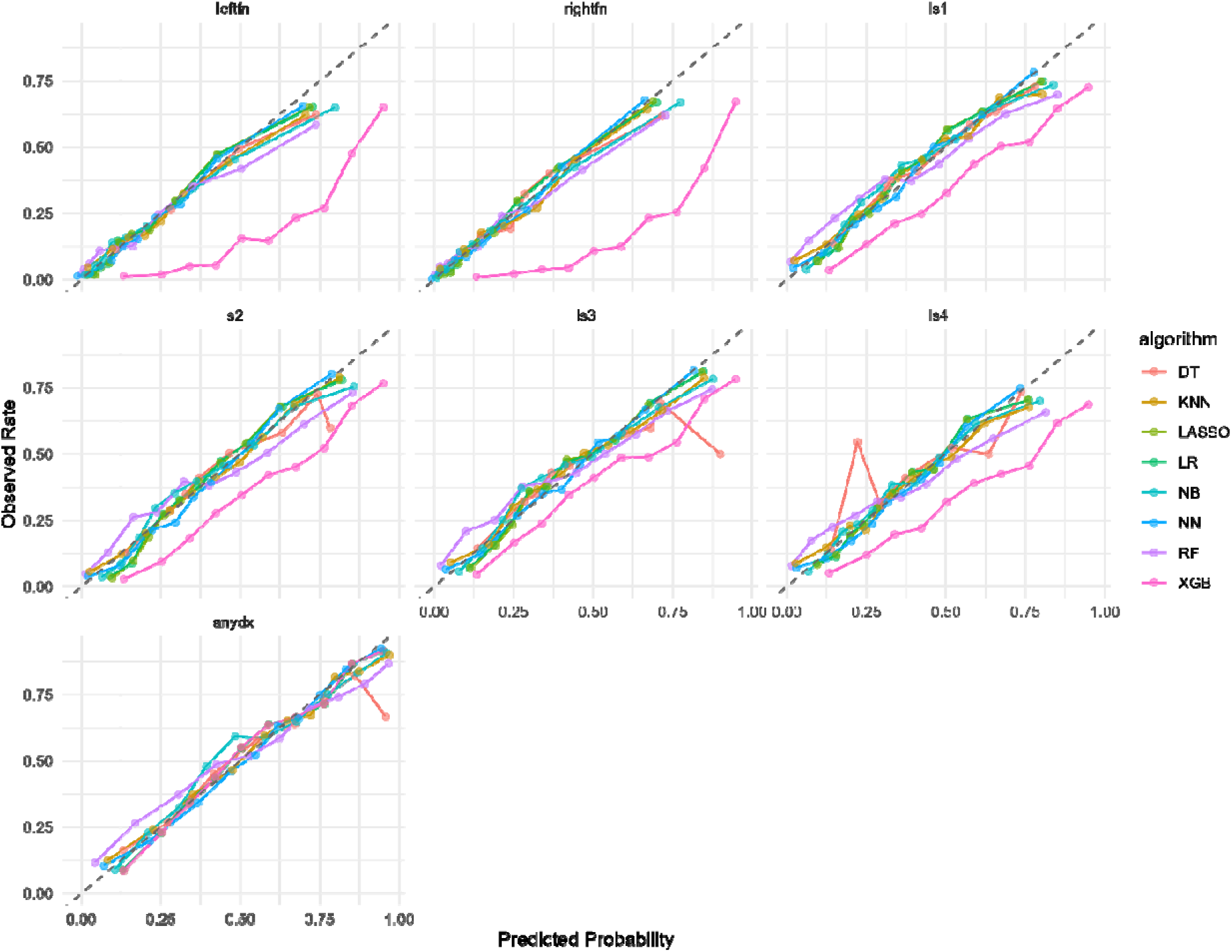
Calibration plots comparing predicted probabilities and observed proportions for eight algorithms across all anatomical sites. FN: femoral neck, LS1: lumbar vertebrae L1, LS2: lumbar vertebrae L2, LS3: lumbar vertebrae L3, LS4: lumbar vertebrae L4 DT: Decision Tree, KNN: K-nearest Neighbors, LR: Logistic Regression, NB: Naïve Bayes, NN: Neural Networks, RF: Random Forest, XGB: XGBoost

## Discussion

Osteoporosis and its consequences represent major public health challenges that necessitate strategies for early diagnosis and intervention at the community level [21]. In this study, we developed self-assessment tools for osteoporosis at multiple anatomical sites – including the left femur, right femur, lumbar vertebrae L1–L4, and any site – for Vietnamese postmenopausal women using a machine learning approach. Eight algorithms were compared, including Logistic Regression, Decision Tree, Random Forest, LASSO, XGBoost, Neural network, Naive Bayes, and K-Nearest Neighboor.

For discrimination, Logistic Regression, LASSO, XGBoost, Naïve Bayes, Neural Network performed equally well due to their ability to balance simplicity, regularization, and computation efficiency. Decision Trees did not perform as good since they are prone to overfitting. K-Nearest Neighbors had an especially low performance compared to other algorithms. This issue is prominently seen in high dimensional data, KNN may to identify meaningful neighbors in low-dimension settings when data has uneven or sparse distribution. Some other possible contributing factors were the sensitivity to feature scaling or the lazy learning characteristic.

We developed an assessment tool and evaluate the predictive performance of the model in various anatomical sites: left femur, right femur, and lumbar vertebrae (L1-L4), and any site. The AUROC of the eight tools indicated a good ability to predict the risk of osteoporosis. Our model outperformed existing screening tools developed for Asian populations, such as those reported by Bui et al., across all anatomical sites except the right femur [7]. Compared with OSTA and OSTC—the two widely used osteoporosis screening tools for Asian postmenopausal women [7], our models showed higher sensitivity, while OSTA tended toward higher specificity. At the left femoral neck, our models achieved 76.0% sensitivity and 74.0% specificity versus OSTA’s 67.3% and 84.6%. In the lumbar spine (L1–L4), the new model maintained greater sensitivity (75–76% vs. 58.4–62.4% sensitivity), whereas OSTA consistently recorded higher specificity (72.9–77.4% vs. 60–64% specificity) [7]. Another more recent study conducted by Bui et al., utilized Random Forest to develop their model. Their model achieved a commendable AUC of 0.86 for predicting outcomes at any anatomical site, demonstrating the strength and potential of machine learning techniques in medical research [8]. However, one of the limitations of their model was that it predicted overall osteoporosis risk rather than site-specific outcomes. Furthermore, the model developed by Bui et al. included numerous laboratory tests as predictors [7], which are typically unavailable in screening settings, thereby limiting its practical implementation. In contrast, our models relied on only four basic predictors suitable for self-assessment, yet achieved comparable discrimination performance. Overall, the new model demonstrates strong potential as a practical screening tool and may represent a preferable alternative for Vietnamese postmenopausal women.

When applying the models or corresponding equations, individuals with scores below the defined threshold are considered at low risk for osteoporosis. For these individuals, routine bone density screening may not be immediately necessary. However, given that the specificity of the tool is not absolute, a low-risk score does not fully exclude the presence of osteoporosis; thus, continued attention to potential symptoms remains important. Conversely, individuals with scores above the threshold are classified as high risk and should undergo further clinical evaluation. Additional diagnostic assessments, such as bone mineral density (BMD) testing, may be warranted to more accurately determine bone health and guide appropriate management strategies based on individual risk profiles. Several limitations of this study should be acknowledged. First, the data were collected from a single hospital in a large urban center in Vietnam, which may limit the generalizability of the findings to postmenopausal women in other regions, particularly rural areas. Second, because of the limited sample size for certain subgroups, we were unable to evaluate model performance within specific high-risk populations, such as individuals with particular lifestyle factors or those using medications known to affect bone mass (e.g., hormonal contraceptives). Finally, this study focused on the development of a new self-assessment tool; therefore, external validation was not performed. Although we reserved a separate dataset for testing, the homogeneity of the data limits the assessment of generalizability. Future studies should include external validation using data from different hospitals or regions to provide more robust evidence of the model’s applicability across diverse populations.

## Data Availability

All data produced in the present study are available upon reasonable request to the authors

## Notes

**Conflict of interest:** The authors have no conflicts of interest to declare for this study

### Competing Interest Statement

The authors have declared no competing interest.

### Funding Statement

This study did not receive any funding

### Author Declarations

The study received ethical approval from the Institutional Review Board of Hanoi Medical University (IRB number: 00003121). The study adhered to ethical principles outlined in the Declaration of Helsinki, ensuring the responsible and ethical use of secondary data

